# Large-scale plasma proteomics in the UK Biobank modestly improves prediction of major cardiovascular events in a population without previous cardiovascular disease

**DOI:** 10.1101/2024.03.13.24304196

**Authors:** Patrick Royer, Elias Björnson, Martin Adiels, Rebecca Josefson, Eva Hagberg, Anders Gummesson, Göran Bergström

**Author notes:** To whom correspondence should be addressed: Prof Göran Bergström, Department of Molecular and Clinical Medicine, Institute of Medicine, Sahlgrenska Academy, Gothenburg University, Gothenburg, Sweden.

## Abstract

**Background and Aims:** Improved identification of individuals at high risk of developing cardiovascular disease would enable targeted interventions and potentially lead to reductions in mortality and morbidity. Our aim was to determine whether use of large-scale proteomics improves prediction of cardiovascular events beyond traditional risk factors (TRFs).

**Methods:** Using proximity extension assays, 2919 plasma proteins were measured in 38 380 participants of the UK Biobank. Both data- and hypothesis-driven feature selection and trained models using extreme gradient boosting machine learning were used to predict risk of major cardiovascular events (MACE: fatal and non-fatal myocardial infarction, stroke and coronary artery revascularisation) during a 10-year follow-up. Area under the curve (AUC) and net reclassification index (NRI) were used to evaluate the additive value of selected protein panels to MACE prediction by Systematic COronary Risk Evaluation 2 (SCORE2) or the 10 TRFs used in SCORE2.

**Results:** SCORE2 and SCORE2 refitted to UK Biobank data predicted MACE with AUCs of 0.740 and 0.749, respectively. Data-driven selection identified 114 proteins of greatest relevance for prediction. Prediction of MACE was not improved by using these proteins alone (AUC of 0.758) but was significantly improved by combining these proteins with SCORE2 or the 10 TRFs (AUC=0.771, p<001, NRI=0.140, and AUC=0.767, p=0.03, NRI 0.053, respectively). Hypothesis-driven protein selection (113 proteins from five previous studies) also improved risk prediction beyond TRFs while a random selection of 114 proteins did not.

**Conclusions:** Large-scale plasma proteomics with data- and hypothesis-driven protein selection modestly improves prediction of future MACE beyond TRFs.

**Structured Graphical Abstract legend.:** 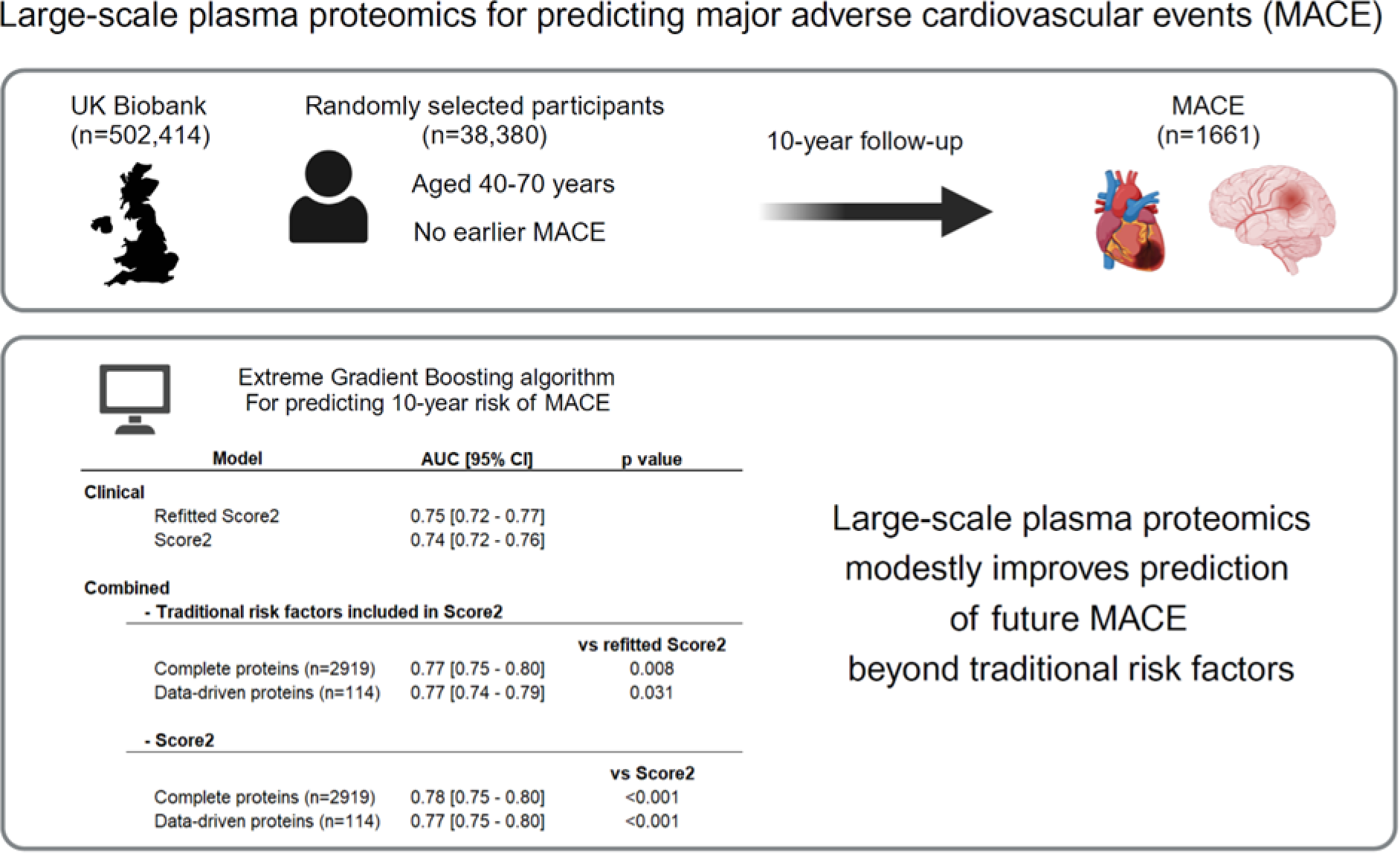

## Introduction

Cardiovascular disease (CVD) is the main global cause of death.^1^ CVD-related morbidity and mortality would likely be reduced if intense primary prevention efforts were focused on the group of people at highest risk. Currently, high-risk individuals are identified by estimating their 10-year risk of major cardiovascular events (MACE) using traditional risk factors (TRFs) ensembled into one of many risk scores.^2, 3^ However, current risk scores lack precision for both the individual and timing of the event,^4, 5^ and there is an intense search for novel biomarkers that can help to improve the scores. The recent development of large-scale, targeted proteomics offers an unprecedented opportunity to test whether novel biomarkers for MACE can be found in the plasma proteome.^6^

Protein sets derived from large-scale proteomics have shown superior prediction when added to clinical risk scores for secondary prevention of MACE in several studies.^7–10^ The results are less clear for primary prevention of MACE. In studies using plasma protein panels derived from aptamer-based affinity reagents, prediction of incident cardiovascular events was equivalent^11^ or modestly superior^12^ to a traditional risk score. However, in a study that used a protein panel derived from paired, nucleotide-labelled antibody probes, prediction of events was clearly superior compared with a model built on TRFs.^13^

In the current study, we tested whether sets of plasma proteins measured using paired antibody probes could predict incident MACE beyond TRFs in a population (38 000 participants from the UK Biobank) without a history of previous cardiovascular disease. Sets of plasma proteins were selected from 2919 measured proteins using both data- and hypothesis-driven techniques.

## Methods

### Study population

The study population comprised participants from the UK Biobank who were randomly selected for plasma proteomic analysis at the baseline visit.^14^ Individuals with earlier MACE and with more than 20% missing protein measurements were excluded. We used ICD-9 and ICD-10 (International Classification of Diseases 10th revision) diagnostic codes and OPCS-4 (OPCS Classification of Interventions and Procedures version 4) when identifying prevalent disease before the base-line examination (Table S1). UK Biobank has approval from the North-West Multi-centre Research Ethics Committee (MREC) as a Research Tissue Bank (RTB). The current study was also approved by the Swedish ethical review authority (2021-04030).

### Definition of outcome data

MACE was defined as fatal or non-fatal cardiovascular events (myocardial infarction, revascularization procedure, ischaemic stroke, and intracerebral haemorrhage) during the 10-year follow-up after inclusion in the study. We used ICD-10 diagnostic codes and OPCS-4 for outcome data (Table S1).

### Traditional risk factors

Baseline characteristics were collected at the baseline examination as previously described.^15^ The 10 TRFs used in the models are: age, sex, systolic blood pressure, current smoking status, diabetes status and age at diagnosis, glycated haemoglobin, estimated glomerular filtration rate, HDL, and total cholesterol.

### Proteomic analyses

Detailed information on the proteomics technology as well as the normalization and quality control steps has already been published.^14, 16^ In brief, 2941 plasma protein analytes corresponding to 2923 unique proteins were measured using the antibody-based Olink Explore 3072 proximity extension assay (PEA) technology. Protein measurements were expressed as normalized protein expression (NPX), a Log2 scale arbitrary unit. In the present study, proteins with more than 20% missing NPX values across samples were excluded and missing values for protein and clinical variables were imputed using the K-nearest neighbours (KNN) algorithm.^17^

### Data analyses and statistics

Extreme gradient boosting machine learning models^18^ were trained with grid search and 5-fold cross-validation to predict the 10-year risk of MACE using different subsets and combinations of clinical and protein data. The data set was randomly divided into a training set (80%) and a test set (20%). Two clinical risk prediction models were chosen as reference: (1) SCORE2 (Systematic COronary Risk Evaluation 2;^19^ SCORE2-Diabetes for individuals with diabetes^20^) was used to calculate the 10-year fatal and non-fatal cardiovascular disease risk for each participant; and (2) a refitted risk score was trained on UK Biobank data using the same TRFs as those included in SCORE2 (termed refitted SCORE2).

Four protein models were tested: a “complete” protein model including all proteins; a “hypothesis-driven” protein model based on proteins found to be predictive of MACE in previous studies;^11–13, 21, 22^ a “data-driven” protein model obtained after a protein feature selection procedure on the training set using the Boruta algorithm;^23^ and a corresponding “random” protein model consisting of random proteins whose number was equivalent to that of the data-driven model. Eight combined models were formed by combining the four protein datasets with either the calculated SCORE2 (one variable) or the TRFs included in SCORE2 (10 variables). The workflow is described in Figure S1.

The performance of the models was assessed using the area under the receiver operating curve (AUC) and the categorical net reclassification index (NRI) using a 5 and 10% risk threshold.^24^ AUC for the SCORE2 and refitted SCORE2 were compared with AUC for the protein and combined models using DeLong’s test.^25^ The calibration of the models was evaluated by plotting reliability curves.

Statistical and machine learning analyses were performed using R version 4.0.4 (R Foundation for Statistical Computing, Vienna, Austria). Baseline characteristics between participants who experienced or did not experience a MACE during the 10-year follow-up were compared using mean, standard deviation and t-test for continuous variables, and proportion and Chi-square test for categorical variables. A two-tailed P value of <0.05 was considered statistically significant.

## Results

### Characteristics of study population

Flowchart of inclusion is presented in Figure S1. In total, 46 799 randomly selected participants from the UK Biobank (total n=502 414) had available data on proteomics and 45 666 of these had no history of MACE at the baseline visit. After excluding individuals and proteins with more than 20% missing protein measurements, the final cohort comprised 38 380 individuals with 2919 unique proteins measured. Baseline characteristics of the participants divided by MACE are presented in Table 1 without imputations. The pattern of missing data is presented in Table S2. Individuals who experienced a MACE during the 10-year follow-up (n=1661, 4.3%) had a much more severe cardiovascular risk factor profile at baseline compared to those who did not experience events (Table 1).

**Table 1.** Baseline characteristics of participants.

|  | Total | MACE within ten years |  |  |
| --- | --- | --- | --- | --- |
|  | n=38380 | No<br>n=36719 (95.7) | Yes<br>n=1661 (4.3) | p value |
| Male sex, n (%) | 17241 (44.9) | 16091 (43.8) | 1150 (69.2) | <0.001 |
| Age (years) | 56.51 (8.12) | 56.31 (8.12) | 60.94 (6.70) | <0.001 |
| BMI (kg/m<sup>2</sup>) | 27.38 (4.77) | 27.33 (4.76) | 28.44 (4.88) | <0.001 |
| Waist (cm) | 90.05 (13.35) | 89.79 (13.29) | 95.81 (13.44) | <0.001 |
| Diabetes, n (%) | 563 (1.5) | 469 (1.3) | 94 (5.7) | <0.001 |
| Age at diabetes diagnosis (years) | 56.60 (7.67) | 56.57 (7.68) | 56.74 (7.68) | 0.836 |
| Systolic blood pressure (mmHg) | 82.28 (10.69) | 82.19 (10.65) | 84.15 (11.24) | <0.001 |
| Diastolic blood pressure (mmHg) | 139.62 (19.55) | 139.29 (19.47) | 146.91 (19.86) | <0.001 |
| HDL cholesterol (mmol/l) | 1.46 (0.38) | 1.46 (0.38) | 1.33 (0.39) | <0.001 |
| LDL cholesterol (mmol/l) | 3.57 (0.87) | 3.57 (0.86) | 3.51 (0.98) | 0.007 |
| Total cholesterol (mmol/l) | 5.72 (1.14) | 5.72 (1.13) | 5.56 (1.29) | <0.001 |
| Triglyceride (mmol/l) | 1.74 (1.03) | 1.73 (1.03) | 1.96 (1.07) | <0.001 |
| eGFR (mL/min/1.73 m<sup>2</sup>) | 94.82 (12.89) | 94.99 (12.77) | 90.88 (14.80) | <0.001 |
| C-reactive protein (mg/l) | 2.59 (4.33) | 2.55 (4.27) | 3.46 (5.50) | <0.001 |
| Glucose (mmol/l) | 5.12 (1.20) | 5.11 (1.16) | 5.48 (1.90) | <0.001 |
| Haemoglobin A1c (mmol/mol) | 36.07 (6.59) | 35.93 (6.32) | 39.28 (10.31) | <0.001 |
| Smoking - current, n (%) | 4117 (10.7) | 3807 (10.4) | 310 (18.7) | <0.001 |
| Smoking - previous, n (%) | 13129 (34.2) | 12486 (34.0) | 643 (38.7) | <0.001 |
| Smoking - never, n (%) | 20968 (54.7) | 20274 (55.3) | 694 (41.8) | <0.001 |
| Hyperlipidemia medication, n (%) | 6085 (15.9) | 5486 (14.9) | 599 (36.1) | <0.001 |
| Hypertension medication,<br>n (%) | 7528 (19.6) | 6872 (18.7) | 656 (39.5) | <0.001 |

MACE, major adverse cardiovascular event; BMI, body mass index; HDL, high-density lipoprotein; LDL, low-density lipoprotein; eGFR, estimated glomerular filtration rate.
Values are expressed as mean and standard deviation for continuous variables, and number and proportion for categorical variables.
t-test and Chi-square test were used for continuous and categorical variables.

### Selection of proteins for prediction models

Missing NPX values represented 2.4% of the total 112 031 220 protein data points and were imputed with the KNN algorithm. Workflow for protein selection is described in Figure S1. In the data-driven protein set, 114 proteins were selected by the Boruta algorithm as being of relevance for MACE prediction after 276 iterations on the training set (n=30 704). In the hypothesis-driven protein set, 113 proteins were compiled from 115 candidate biomarker proteins identified in five previous publications;^11–13, 21, 22^ the remaining two candidate biomarkers identified in these publications (AGP1 and TREM1) had not been analysed in the current UK Biobank sample. The random protein set was created with the same number of proteins (114) as in the data-driven protein set. The complete protein set included all 2919 proteins. The data-driven and hypothesis-driven protein sets had 20 proteins in common while the random protein set had two and four proteins in common with the data-driven and hypothesis-driven protein sets respectively. A list of all proteins measured, and the different protein sets is found in Table S3.

### Performance of prediction models

The predictive performance of the clinical, protein and combined models was evaluated on the test set (n=7676) and AUC results are presented in Table 2. SCORE2 and refitted SCORE2 predicted MACE with an AUC of 0.740 and 0.749 respectively. The complete, data-driven, and hypothesis-driven protein models all had numerically higher AUC (0.773, 0.758, 0.759, respectively) but the difference was only significant for the complete protein model (p=0.003 and 0.014 compared to SCORE2 and refitted SCORE2, respectively). The random protein model performed significantly worse than the two clinical models (AUC=0.712).

**Table 2.**
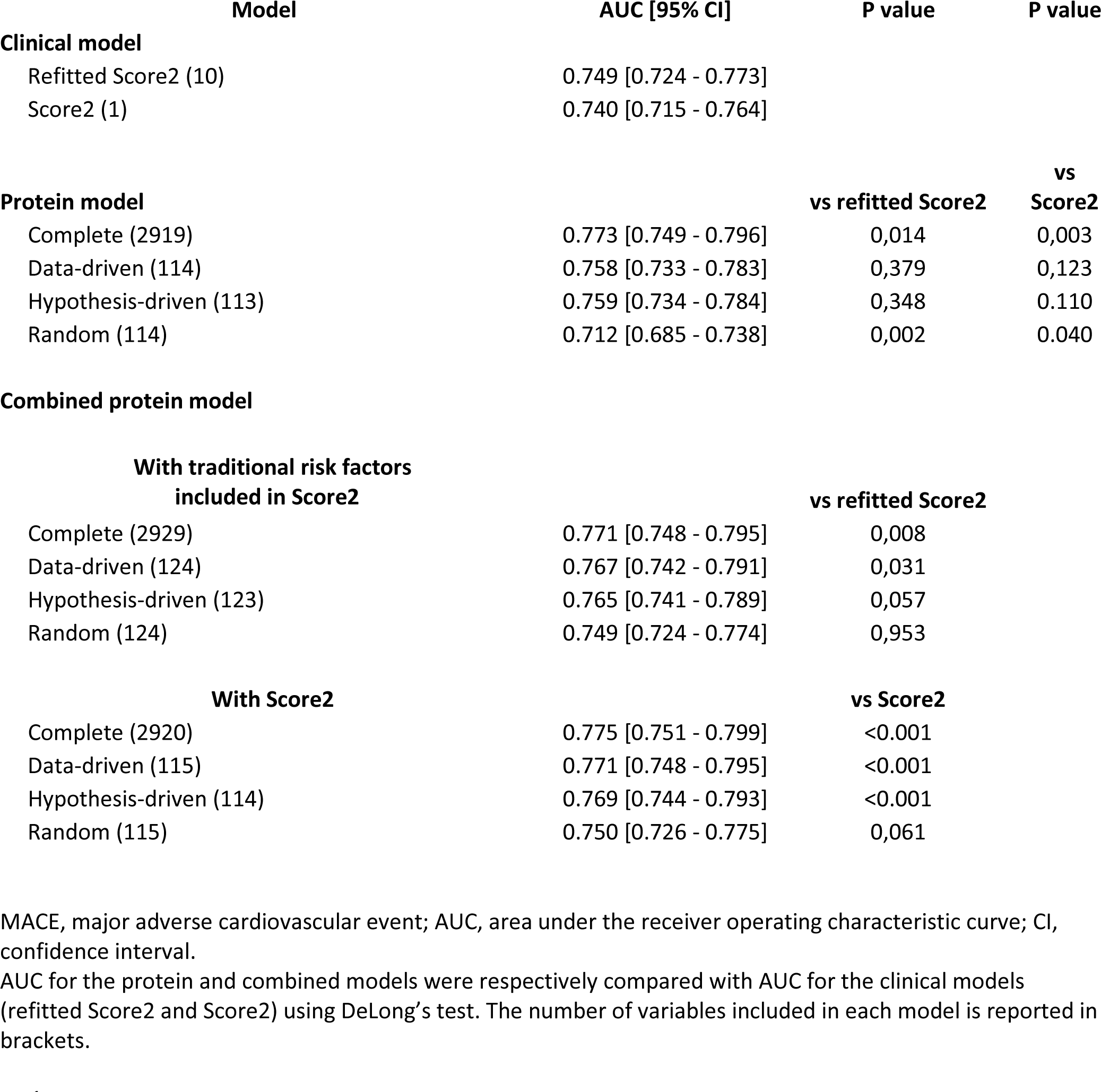
Performance of the clinical, protein and combined models for predicting the 10-year risk of MACE.

When the proteins selected in the data-driven model were combined with the 10 TRFs used in SCORE2, they significantly outperformed predictions compared to both SCORE2 (ΔAUC = +0.029, p<0.001) and refitted SCORE2 (ΔAUC = +0.016, p=0.031). When the proteins selected in the hypothesis-driven model were combined with TRFs, they numerically increased the AUC compared to refitted SCORE2 (ΔAUC = +0.018, p=0.057). The hypothesis-driven protein set significantly increased AUC when combined with SCORE2 compared to SCORE2 alone (ΔAUC = +0.029, p<0.001). The complete protein set did not outperform the data-driven protein set when combined with either TRFs (p=0.198) or SCORE2 (p=0.310). The random protein set combined with TRFs or SCORE2 did not significantly change the discrimination compared to the clinical models alone.

Reclassification tables and the calculated NRI for all combined models are presented in Figure 1 and Figure S2. At the 5% risk threshold, NRI ranged from 0.026 (random protein set) to 0.044 (complete protein set), and no protein set significantly improved the reclassification when combined with TRFs and compared with the refitted SCORE2 (Table S2). However, at the 10% risk threshold, the NRI was significantly increased when the complete protein set (NRI=0.046, p=0.039) or the data-driven protein set (NRI=0.053, p=0.020) was combined with TRFs and compared with the refitted SCORE2 (Figure 1B and D). A significantly increased NRI was also seen when the data-driven (0.048, p=0.032) or the hypothesis-driven (0.049, p=0.046) protein sets were combined with SCORE2 at the 5% risk threshold. Further, all four protein sets significantly increased NRI when combined with SCORE2 at the 10% risk threshold (0.112, 0.140, 1.148 and 0.061, p<0.01, for the complete, data-driven, hypothesis-driven, and random protein sets, respectively).

**Figure 1.**
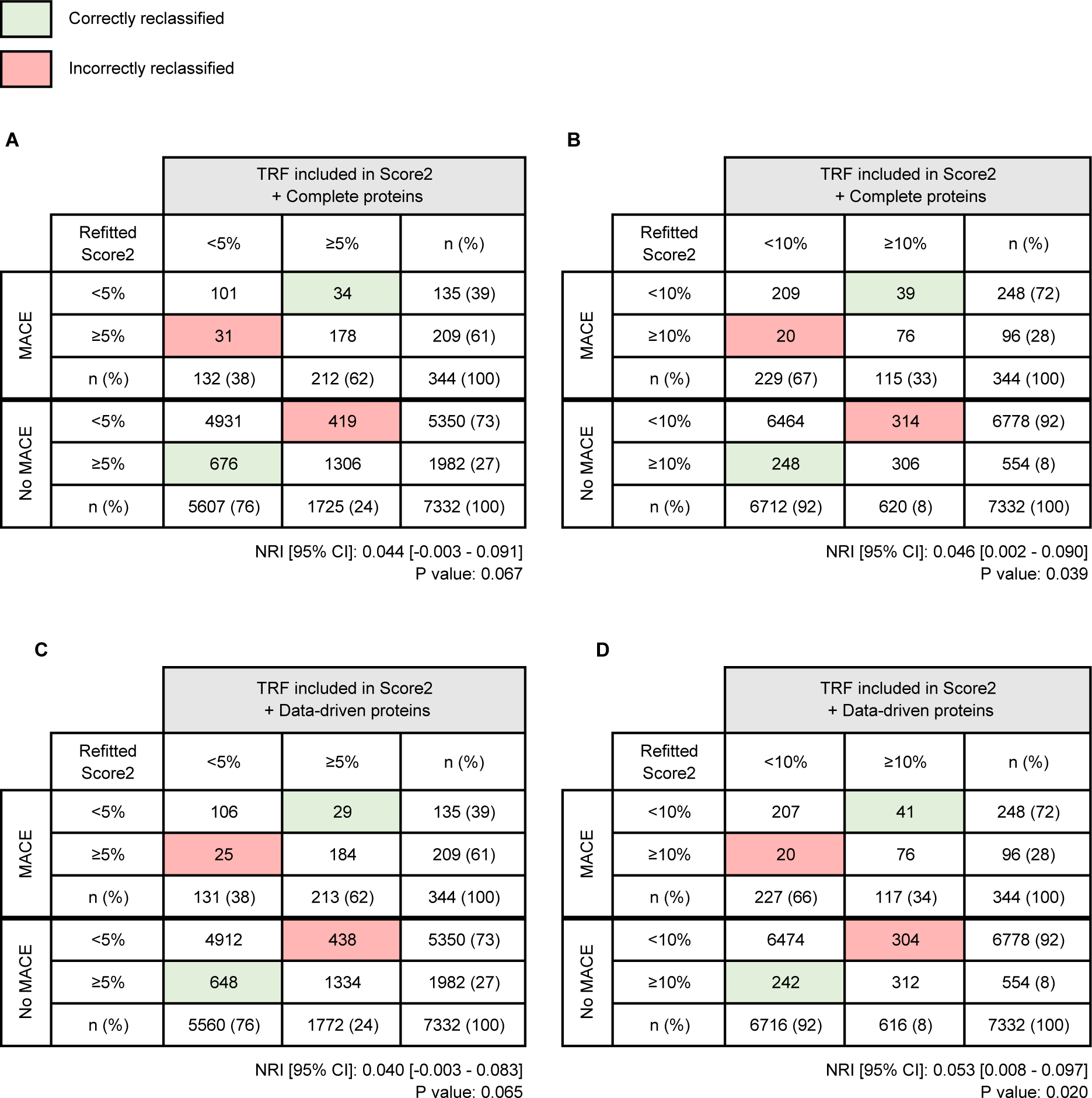
Reclassification tables. The figure shows reclassification results in the test set (7676 participants and 344 events) when (**A,B**) complete (2919) and (**C,D**) data-driven (114) protein sets are combined with the 10 TRFs included in SCORE2 for predicting the 10-year risk of first major cardiovascular event (MACE). Panels A and C show net reclassification index (NRI) for the 5% risk threshold, and panels B and D show NRI for the 10% risk threshold. CI, confidence interval.

Model performance at the 5% and 10% cut-off values for 10-year risk of MACE can be analysed in detail using the reclassification tables (Figure 1 and Figure S2). Using the 5% cut-off, the models tend to improve classification mainly in the non-MACE group. On the contrary, when using the 10% cut-off, classification is mainly improved in the MACE group. For example, the largest reclassification in the non-MACE group was seen when the complete protein set was combined with SCORE2 and compared to the SCORE2 model using a 5% cut-off: 1290 individuals were correctly reclassified while only 187 were incorrectly reclassified (15% correct reclassification). The largest reclassification in the MACE group was seen when the hypothesis-driven protein set was combined with SCORE2 and compared to the SCORE2 model using the 10% cut-off for risk. A total of 73 individuals were correctly reclassified while only 10 were incorrectly reclassified (18% correct reclassification).

As shown by reliability curves in Table S4, the clinical, protein and combined models were correctly calibrated.

The 113 candidate protein biomarkers presented in five previous studies in primary prevention^11–13, 21, 22^ and measured in the UK Biobank are shown in Table S4. Three proteins (GDF15, MMP12 and NTproBNP) were found in 3 studies, ten proteins were found in 2 studies while the remaining 111 proteins were found only once.

## Discussion

In this study, we used data from individuals without previous CVD from the UK Biobank to test whether prediction models for MACE could be improved by addition of subsets derived from 2919 measured proteins. Using a data-driven feature selection, we identified 114 proteins as being of relevance for prediction of MACE. Prediction by this panel of proteins was equal to that of SCORE2 and a refitted model based on the TRFs used in SCORE2 but trained on UK Biobank data. More importantly, the discriminative capacity was significantly increased when the 114 proteins were used in combination with the TRFs used in SCORE2 (10 variables) or the SCORE2-calculated risk (one variable). We also showed that a hypothesis-driven dataset of 113 proteins previously suggested as biomarkers of CVD^11–13, 21, 22^ added discriminative capacity to SCORE2. A model using 114 randomly selected proteins did not improve discrimination, supporting the specificity of the selected proteins. We consider our protein selection successful since the AUC of the combined complete protein model was not different from the AUC of the combined model using data-driven protein selection.

There are a few large studies in the literature using targeted proteomics to predict CVD events in populations without previous CVD^11–13, 21, 22^. In a study that used aptamer technology and a case-control design, a panel of 13 proteins was shown to be equally effective to a traditional risk model in predicting CVD.^11^ In another study using paired antibody probes in a case-control design, a model based on 50 proteins was superior to a model using refitted TRFs.^13^ Our findings support and extend this result by showing that a data-driven selection of proteins, from a large set of proteins measured using paired antibody probes, can be used to improve classification of MACE in a large unselected population sample. A recent study using aptamer technology showed that 70 proteins in combination with TRFs significantly improved the AUC slightly but only increased NRI significantly when a high risk threshold was used.^12^ There are also two studies showing associations of protein subsets with CVD independent of TRFs.^21, 22^ In the above five studies, a total of 115 unique proteins has been suggested as candidate biomarkers. Our study supports the selection of these candidate proteins since our hypothesis-driven panel of proteins also added discriminatory capacity for MACE beyond that of SCORE2.

A few of the individual protein candidate biomarkers from our own study and the five previously published papers^11–13, 21, 22^ are common to more than one study (e.g., GDF15, MMP12 and NTproBNP are in three of the previous studies and in the current study), but most candidate proteins are unique to one study. This lack of reproducibility could indicate that the biological signal of CVD risk is not strong enough to overcome variation in data design, cohort definitions and choice of outcome. It is also possible that the field of large-scale proteomics is not mature enough to provide stable measurements of large series of biomarkers under different conditions. Continued work on analytic validity, repeatability, replication, and external validation is required to improve candidate biomarker generalizability.^26^

An important question to address is whether the significant shifts in discriminatory capacity we found are of clinical benefit. The shifts in AUC were numerically small (up to 0.035) which was also true for the improvements in NRI (0.04-0.15). Depending on the risk threshold used and the model tested, up to 18% of participants in the group that will later suffer MACE could be correctly reclassified by combining the data-driven selection of protein biomarkers with SCORE2. This number are close to the ones presented recently when evaluating large-scale proteomics for prediction of CVD events in a large Icelandic population^12^ and appear to represent a modest improvement. It also appears from our and this recent analysis^12^ that the benefits of adding proteins to a risk prediction model is best seen at a higher risk threshold. If true, proteins could be better used as a diagnostic test in a population with a high-pretest likelihood of diseases than as a screening tool in the general population at low risk. This notion is also supported by the success of protein biomarkers in secondary prevention^7–10^ relative to primary prevention.^11–13, 21, 22^

Our study has several limitations. First, our study lacks an external validation set, which limits the generalisability of our findings. Second, proteins were measured using dual binding affinity proteomics; although this method is known to have high protein target specificity and a high number of phenotypic associations,^27^ we cannot be sure that our findings can be replicated using other proteomic platforms. Third, this study was not designed to identify individual biomarkers, but to create a reliable panel of proteins that could predict MACE; we did not test for a potential causal role of individual proteins in the development of MACE since this was not the focus of this paper. Fourth, this report describes some of the potential benefits of using protein scores for risk estimation. A detailed health-economic assessment of both costs and benefits of using large-scale proteomics needs to be performed to assess the net clinical benefit^28^ of these improvements, which is beyond the scope of this report. Further, intervention trials are needed to test whether the incremental improvement in classification of risk, potentially introduced by proteomics, can be transferred into fewer CVD events.

## Conclusion

Using machine learning and a large set of proteins measured using dual antibody probes, we could improve identification of MACE with a panel of 114 proteins selected using a data-driven technique. Similar, although not as convincing, improvements were achieved using a hypothesis-driven panel of 113 proteins. The improvements are, however, relatively small and the clinical utility of adding these biomarkers in primary prevention will have to be established.

## Acknowledgements

This research has been conducted using the UK Biobank Resource under Application Number 82018. The authors are very grateful for the excellent editorial assistance of Rosie Perkins.

## Funding

This study was supported by the Swedish Heart Lung Foundation (20210383), the Swedish Research Council (2019-01140) and grants from the Swedish state under the agreement between the Swedish government and the county councils, the ALF-agreement (ALFGBG-718851, ALFGBG-991828).

## Disclosure of interest

No author reports conflicts of interest.

## Data availability statement

The data that support the findings of this study are available from UK Biobank.

## Key Question

Can plasma proteins predict the 10-year risk of first major adverse cardiovascular events (MACE)? Can they outperform or add a prognostic value to clinical models?

## Key Finding

An extreme gradient boosting model trained and tested on 30,704 and 7,676 adults from the UK Biobank with a data-driven set of 114 plasma proteins improved the prediction of first MACE when combined to clinical models Score2 and a refitted version of Score2. A protein model alone including a large set of 2919 plasma proteins outperformed the predictions of the clinical models.

## Take-home Message

Plasma proteomics may improve the clinical predictions of first MACE. Further research should precise the cost benefits and the optimal size of the predictive protein panel to use.

## Notes

### Competing Interest Statement

The authors have declared no competing interest.

### Author Declarations

UK Biobank has approval from the North-West Multi-centre Research Ethics Committee (MREC) as a Research Tissue Bank (RTB). The current study was also approved by the Swedish ethical review authority (2021-04030).

